# Evaluation of Provider Clinical Decision Support System Adoption Rates by Patient Race and Sex

**DOI:** 10.64898/2026.07.21.26358637

**Authors:** Ratnalekha V. N. Viswanadham, Simon A. Jones, Kellie Owens, Safiya I. Richardson

## Abstract

**BACKGROUND:** Clinical decision support (CDS) systems can improve care quality, but their implications for equity remain uncertain. We examined whether provider response to CDS alerts differed by patient race and sex in primary care, and whether differences in alert exposure helped explain any observed variation.

**METHODS AND PRINCIPAL FINDINGS:** We conducted a retrospective study using EHR data from a New York City academic health system, focusing on alert-based CDS during outpatient primary care. Logistic regression was used to estimate the likelihood of alert engagement by patient race and sex, while adjusting for encounter and provider factors. We used a generalized structural equation model to assess mediation by alert type, decomposing direct and indirect effects of demographics on response.

Direct effects suggest that providers may respond differently to alerts based on patient identity, consistent with interpersonal bias, in which implicit or explicit attitudes shape clinical behavior, and on the context of the visit. Indirect effects highlight disparities in how alerts are assigned across groups, indicating that algorithmic or systemic bias may be embedded within the technology itself. Estimated mediated pathways suggest that even when providers respond uniformly to alerts, unequal exposure can still produce inequitable outcomes.

**DISCUSSION:** The findings highlight that the type of CDS triggered plays a significant role in differential CDS responses, with provider- and patient-related factors evident in these differences. These findings underscore the need to evaluate not only provider behavior but also the logic and distribution of CDS tools themselves, as both can contribute to disparities in care delivery. Further research should also focus on looking for the potential health impact of the differential response.

**AUTHOR SUMMARY:** Digital tools meant to standardize care can unintentionally contribute to which patients receiving care. We investigate whether providers’ use of these tools is related to a patient’s identity or influenced by the types of tools provided to them in primary care. Using electronic health record data from a large urban health system, we find that providers’ responses to alerts are shaped not only by patient identity but also by the nature of the alert itself. Direct effects suggest that providers may engage differently with CDS based on patient demographics, indicating potential interpersonal bias. Indirect effects reveal that certain patient groups are more or less likely to receive specific types of alerts, indicating embedded algorithmic or systemic bias. These findings underscore the importance of evaluating both provider behavior and the design of CDS tools when assessing equity in digital health. Even when providers respond consistently, unequal exposure to alerts can produce inequitable outcomes. Our results underscore the need for more transparent and equity-aware CDS design and implementation strategies that consider both human and technological sources of bias.

## INTRODUCTION

Clinical decision support (CDS) systems embedded in electronic health records (EHRs) can improve the quality and consistency of care. (1, 2) These systems are often integrated into routine workflows through alerts that appear during prescribing, ordering, or documentation and prompt guideline-concordant actions. Prior studies and meta-analyses have shown that CDS can improve adherence to evidence-based treatment and preventive care and may reduce morbidity, adverse events, and diagnostic delays. (3–6) In primary care, CDS has been associated with improvements in antibiotic prescribing, cancer screening, and cardiovascular risk management. (7–10)

However, the benefits of CDS are not necessarily distributed evenly across patient populations. CDS can reduce unwarranted variation in care, but it may also reflect underlying differences in clinical workflow, documentation practices, alert design, or model performance that affect how often patients receive alerts and how often clinicians act on them. (11–14) Prior work has shown that inequities in CDS performance on provider engagement can arise from biased training data, assumptions in decision rules, or differences in workflow across settings and patient groups. (15) In addition, clinicians may engage differently with CDS depending on patient context, time pressure, implicit bias, or the perceived relevance of the recommendation. (16, 17)

Race and sex are two important axes along which inequities in healthcare are often observed. Both have been linked to differences in access, treatment patterns, and outcomes, and they may also shape how decision support is delivered and used in clinical practice. (18) Yet most CDS equity studies have focused on either race or sex alone, rather than their combined relationship to alert exposure and alert response. A recent scoping review found that relatively few CDS studies explicitly evaluated equity impacts, underscoring a gap in how digital tools are assessed in practice. (19)

Primary care is a useful setting for examining these questions because it is a common point of contact for prevention, chronic disease management, and acute care. Small differences in how alerts are generated or acted upon may accumulate over time into meaningful differences in care delivery. To address this gap, we examined whether provider responses to CDS alerts differed by patient race and sex in a large primary care EHR dataset, and whether any observed differences persisted after accounting for patient, encounter, provider, and alert-level characteristics. We also assessed whether alert type helped explain differences in alert response, with the goal of separating patterns of alert exposure from patterns of provider action. This study adds to the literature on equity in digital health by providing a real-world assessment of CDS use in a diverse clinical setting.

## RESULTS

### Study Population and Descriptive Data

We analyzed a total of 2,849,606 alerts fired across 258,254 unique patients and 512 providers, encompassing 96 unique alerts over 448,211 outpatient encounters over two one-year periods: July 1, 2018–June 30, 2019 (pre-COVID) and July 1, 2021–June 30, 2022 (post-COVID). Patients ranged in age from 18 to 105 years, with a mean age of 54.83 (SD 17.05) and a median age of 56.

The racial distribution was 61.95% Non-Hispanic/Latinx White (“White”), 12.37% Non-Hispanic/Latinx Black/African American (“Black”), 6.21% Asian, and 4.06% Hispanic/Latinx, with other categorizations – Native American/Alaska Native (NAAN), Native Hawaiian/Pacific Islander (NHPI), Non-Hispanic/Latinx Multiple Races/Ethnicities (“Multi”), and not included in the U.S. Census tabulations (“Other”) – had small cell counts. We did not bin these results due to the limited research on these races and ethnicities in health technology, such as telemedicine. (20) Across racial/ethnic groups, White patients represented the majority of visits in all categories. For example, they accounted for 67.0% of Follow-Up Care visits and 60.2% of Preventive Care visits.

In contrast, Asian patients had a comparatively higher proportion of Preventive Care visits (33.6% of all Asian visits), and Hispanic/Latinx patients had a relatively higher share of Acute/Sick Care visits (5.2% of Hispanic visits vs. 1.7% overall). New Patient Evaluations were proportionally higher among patients categorized as (14.3%). Patients identifying with multiple races had a slightly higher proportion of Preventive Care visits than the total sample (30.1% vs. 25.3%).

Overall, Follow-Up Care visits were the most common, accounting for 64.4% of all visits, followed by Preventive Care and Wellness (25.3%) and New Patient Evaluations (8.7%). Visits for Acute or Sick Care were less frequent (1.7%), and Other visits (e.g., nurse visits) were rare (<0.1%). Therefore, “Other” visits were excluded from analyses. Provider roles included attending physicians (88.09%), nurse practitioners (7.03%), and physician assistants (4.88%). However, the provider type was removed from the regressions due to collinearity and a lack of convergence.

### Outcome Data

A total of 96 distinct alert types fired times during the study period. Alerts were grouped into clinical categories: preventive care (the majority of alerts fired), chronic disease management, vaccination (the smallest category but the third most alerts fired), screening, medication stewardship (the majority of alerts categorized), and social determinants of health (the fewest alerts fired). Overall, 14.65% of alerts resulted in provider action. The majority of alerts were shown during work hours between 8 AM and 6 PM (97.08%), so, for regressions, an interaction variable was created between the continuous variable of the hour an alert was fired and whether the alert was shown during work hours to understand the association between an alert being shown later in the day and whether a provider responds to it. The majority of alerts occurred during weekdays and were triggered after the acute COVID-19 pandemic (75.95%). A provider saw an average of 6 to 7 alerts per encounter (mean 6.48 alerts).

Descriptive statistics are presented in Table 1.

**Table 1:**
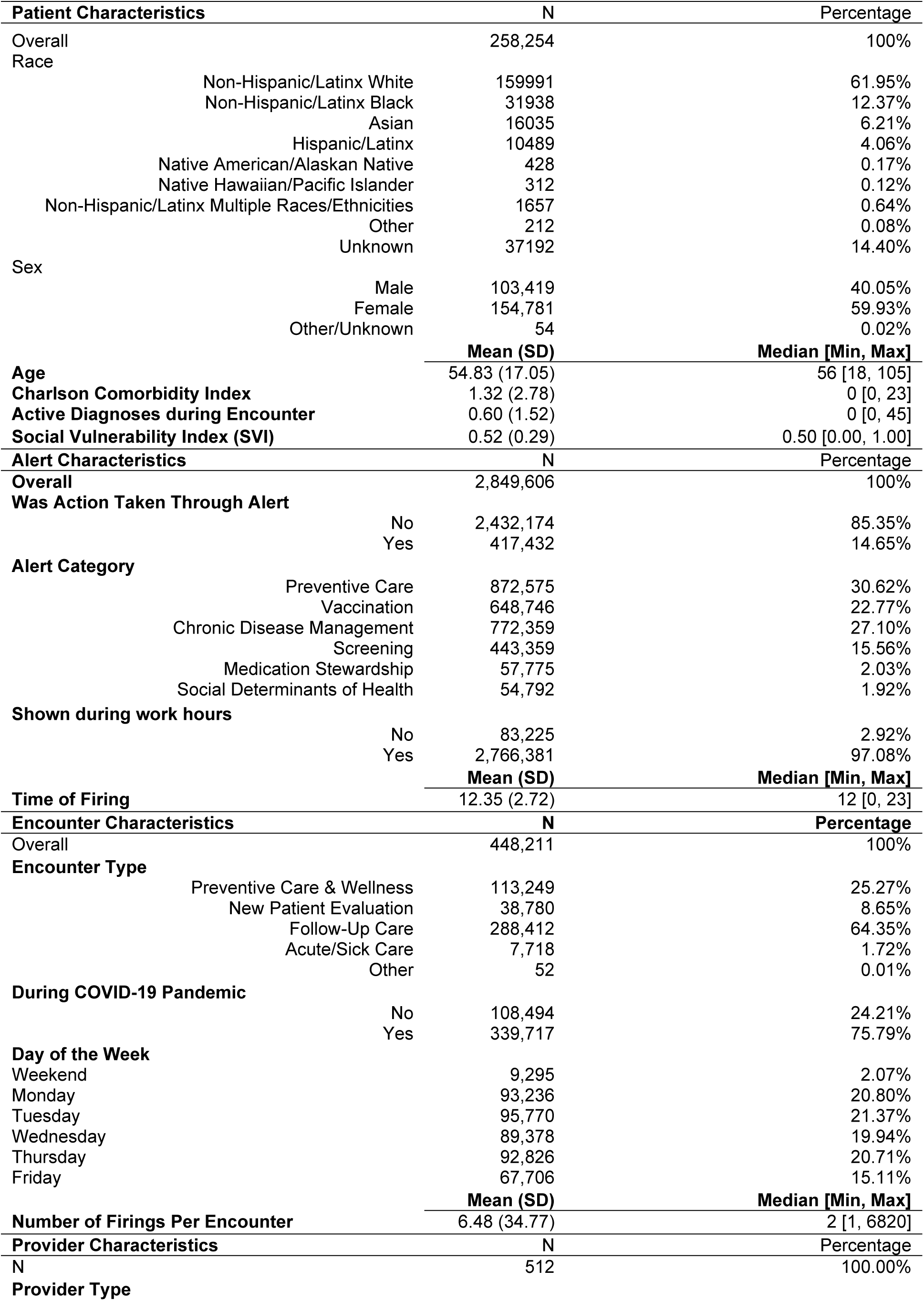
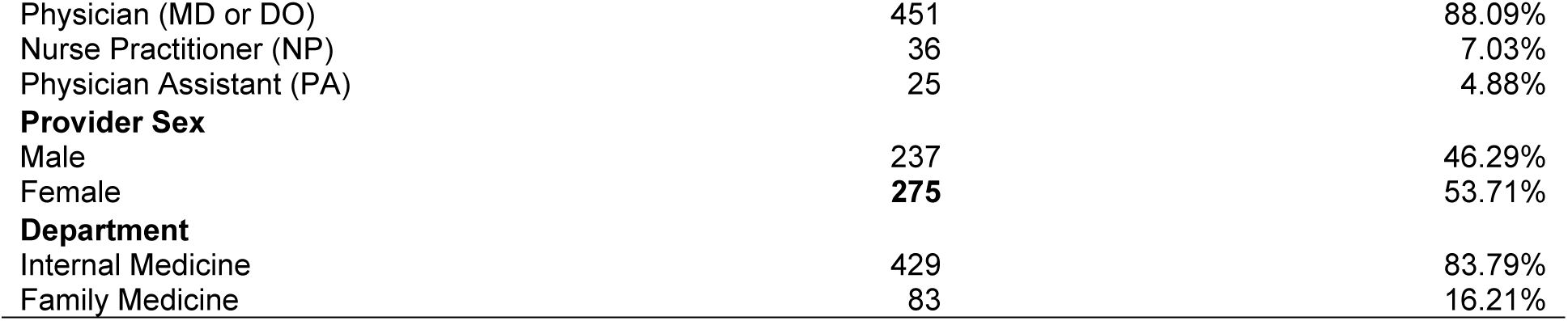
Descriptive Statistics of Patients, Alerts, Encounters, and Providers.

### Logistic Regression

We assessed whether including patients with unknown race/ethnicity or with “other”/unknown sex would affect coefficients in controlled regressions. The exclusion did not significantly change the model fit or coefficients. Therefore, for interpretability, we excluded patients for whom race/ethnicity was unknown, or sex was classified as “other” or unknown. We also compared model fits with and without alert fixed effects and clustering on provider ID and determined that both should be included. The coefficient comparisons can be found in S1 Table 1.

Adjusting for the alert ID and the provider reveals that context factors are significantly correlated with whether an alert has been acted upon. No patient race and ethnicity variables showed a statistically significant relationship to action taken through the alert. Follow-up care visits had fewer alerts acted upon by providers (β = -0.331, ARR = 0.718, p < 0.001), as did acute care visits (β = -0.384, ARR = 0.681, p < 0.001). Alerts fired during the COVID-19 period were significantly less likely to result in action (β = −0.246, ARR = 0.78, p < 0.001), suggesting pandemic-related disruptions in CDS engagement. Alerts later in the day during work hours are slightly less likely to be acted upon, suggesting workflow congestion or alert overload (β = −0.028, ARR = 0.972, p < 0.001).

When not controlling for the alert and provider, the logistic regression revealed significant variation in alert response rates by patient race, while maintaining the significance of context variables. Notably, compared to White patients, Non-Hispanic/Latinx Black patients had a slightly lower response to alerts (β = -0.0177, AR = 0.9825, p < 0.001), as well as patients of multiple Non-Hispanic/Latinx races and ethnicities (β = -0.0130, AR = 0.9871, p < 0.05). In contrast, patients who identified as Asian, Hispanic/Latinx, NA/AN, NH/P, and “Other” had significantly more responses to alerts compared to White patients. Female patients had a reduced response to alerts compared with male patients (β = -0.0628, ARR = 0.9391, p < 0.001). Female providers responded less to alerts than male providers (β = -0.0198, ARR = 0.9804, p < 0.001), and providers in family medicine departments were more likely to respond to alerts than those in internal medicine (β = 0.3689, ARR = 1.4461, p < 0.001).

Notably, the clinical category of the alert significantly correlated with whether providers responded to them. Compared to Preventive Care alerts, providers responded significantly less to alerts about Chronic Disease Management, Vaccination, Screening, and Social Determinants of Health. Medication Stewardship was not significantly associated with a provider taking action on the alert.

Figure 1 and Figure 2 illustrate the odds ratios for the adjusted (1) and unadjusted (2) logistic regression models. Raw coefficients and adjusted odds ratios are available in S1 Table 2 (adjusted logistic regression) and S1 Table 3 (unadjusted logistic regression).

**Figure 1:**
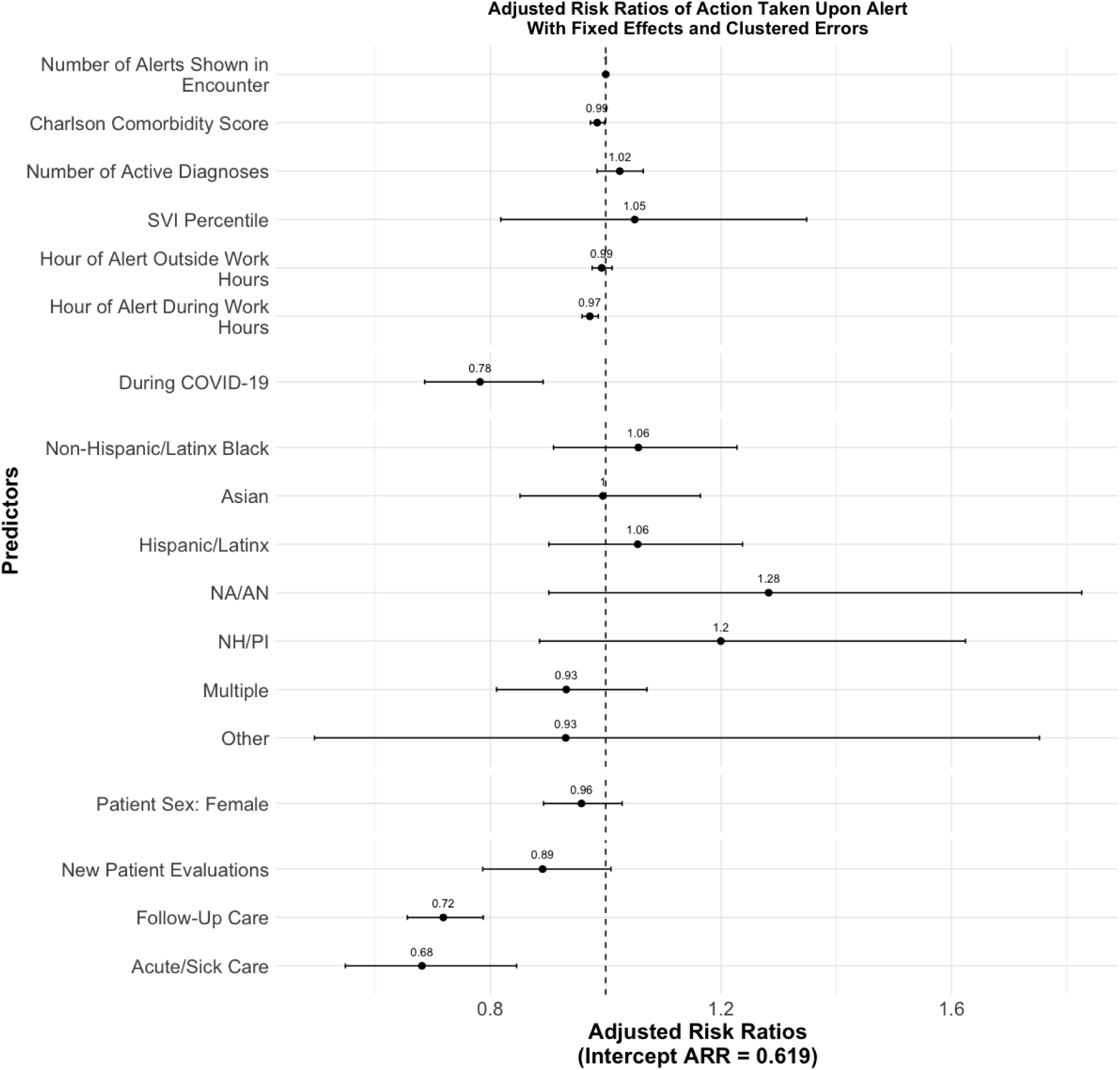
Logistic regression of whether a provider took action through an alert against patient and visit characteristics. This regression adjusts for alert fixed effects and clusters errors on provider ID.

**Figure 2:**
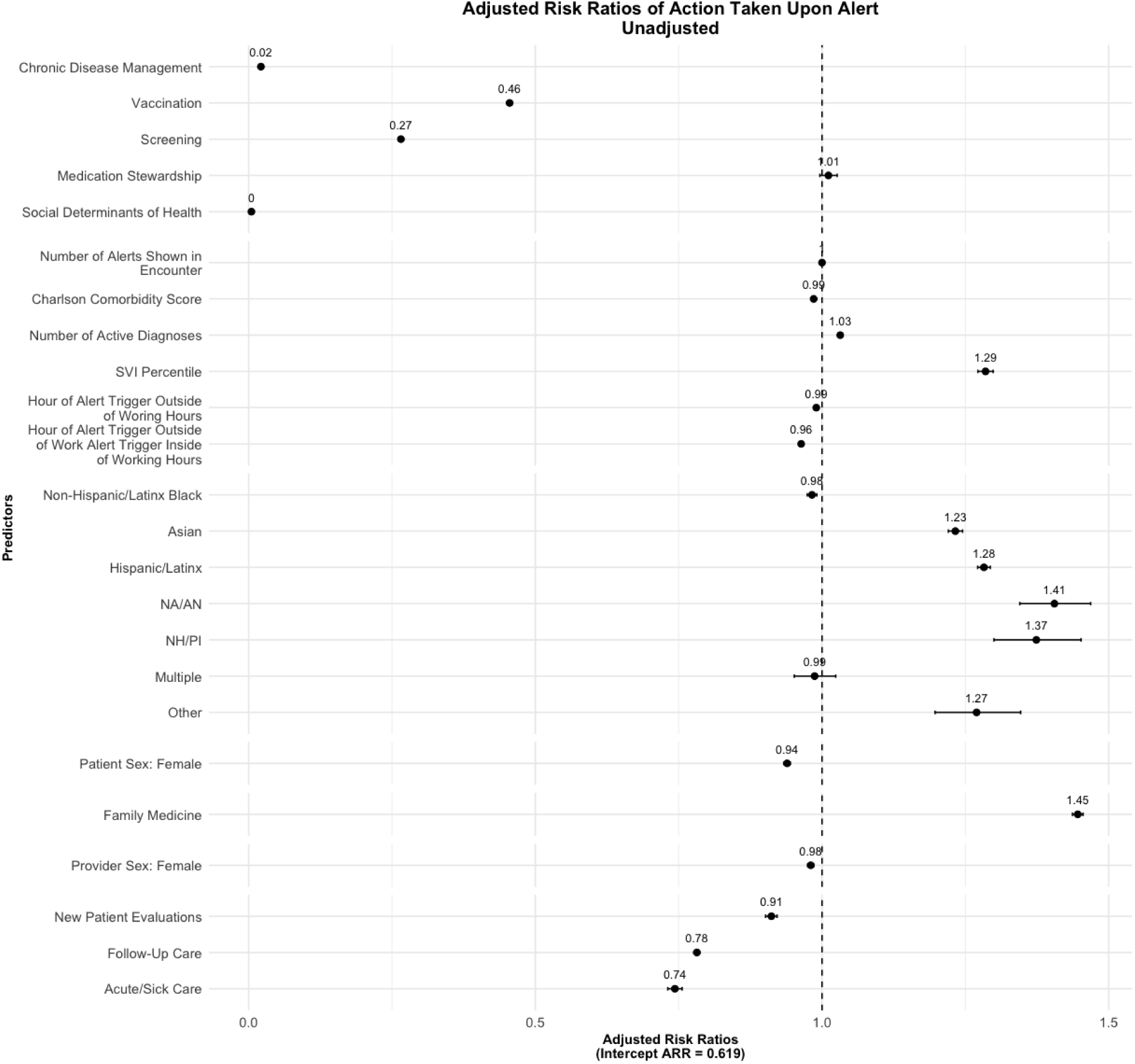
Logistic regression of whether a provider took action through an alert against patient and visit characteristics. This regression does not adjust for alert fixed effects or cluster errors at the provider ID level.

### Multinomial Logistic Regression

Given that the alert category was significantly correlated with a provider’s response to the alert, the goal of the multinomial regression was to evaluate whether patient characteristics were associated with the presence of a specific alert category during an encounter. We estimated a multinomial logit model to examine predictors of the type of clinical decision support (CDS) alert that was fired, with Preventive Care and Wellness alerts as the reference category. Relative risk ratios (RRRs) and 95% confidence intervals (CIs) were calculated for each alert type, adjusting for patient demographics, visit characteristics, comorbidity burden, social vulnerability, and provider department.

Several patient characteristics were significantly associated with increased likelihood of receiving Chronic Disease Management alerts. Compared to Non-Hispanic/Latinx White patients, Non-Hispanic/Latinx Black (RRR = 1.31, 95% CI [1.30, 1.33]), Hispanic/Latinx (RRR = 1.10, CI [1.09, 1.12]), NA/AN (RRR = 1.08, CI [1.00, 1.16]), Multiple race/ethnicities (RRR = 1.17, CI [1.12, 1.22]), and Other (RRR = 1.15, CI [1.06, 1.26]) patients were significantly more likely to receive these alerts. Asian (RRR = 0.84, CI [0.83, 0.86]) and NH/PI (RRR = 0.49, CI [0.44, 0.53]) patients were significantly less likely. Female patients had higher odds of receiving Chronic Disease alerts (RRR = 1.30, CI [1.29, 1.31]). New patient evaluations (RRR = 1.15) and follow-up care (RRR = 1.38) were associated with increased alert firing, whereas acute/sick care visits did not show significant differences. Higher Charlson Comorbidity Scores were strongly associated with increased alert firing (RRR = 1.15), while more active diagnoses were associated with reduced odds (RRR = 0.95). Higher SVI percentiles were associated with lower odds of receiving these alerts (RRR = 0.69).

Vaccination alerts were less likely to fire for most racial/ethnic minority groups. Hispanic/Latinx (RRR = 0.70), NA/AN (RRR = 0.58), Multiple (RRR = 0.60), and Asian (RRR = 0.86) patients had significantly lower odds of receiving vaccination alerts compared to Non-Hispanic/Latinx White patients. Female patients were more likely to receive vaccination alerts (RRR = 1.27). Follow-up (RRR = 1.39) and acute/sick care visits (RRR = 1.32) were associated with increased alert firing. Higher comorbidity scores increased odds (RRR = 1.05), while higher SVI percentiles decreased odds (RRR = 0.78).

Screening alerts showed pronounced disparities. Asian patients had the highest increased odds (RRR = 1.63), followed by Other (RRR = 4.32), NH/PI (RRR = 1.19), NA/AN (RRR = 1.16), and Non-Hispanic/Latinx Black patients (RRR = 1.14). Hispanic/Latinx patients had reduced odds (RRR = 0.89). Female patients were dramatically more likely to receive screening alerts (RRR = 12.10). New patient evaluations increased odds (RRR = 1.27), while follow-up and acute/sick care visits decreased them. Higher SVI percentiles were associated with reduced odds (RRR = 0.75).

Medication alerts were significantly less likely to fire for all racial/ethnic minority groups. NH/PI (RRR = 0.04), NA/AN (RRR = 0.27), Hispanic/Latinx (RRR = 0.38), Non-Hispanic/Latinx Black (RRR = 0.45), and Other (RRR = 0.25) patients all had substantially reduced odds compared to Non-Hispanic/Latinx White patients. Female patients were more likely to receive these alerts (RRR = 1.98). Follow-up care (RRR = 4.48) and acute/sick visits (RRR = 2.66) were strongly associated with alert firing. Higher SVI was associated with increased odds (RRR = 3.59) compared with other alert types.

Social Determinants of Health (SDoH) alerts showed the most extreme disparities. Asian (RRR = 3.27), NA/AN (RRR = 3.39), Non-Hispanic/Latinx Black (RRR = 2.48), and Other (RRR = 9.80) patients had dramatically higher odds of receiving these alerts. NH/PI patients had near-zero odds (RRR ≈ 0). Female patients (RRR = 2.94) and new patient evaluations (RRR = 2.42) were strongly associated with the firing of SDoH alerts. Higher SVI was associated with reduced odds (RRR = 0.30), suggesting that alerts may not be targeting the most socially vulnerable patients.

This model reveals substantial disparities in CDS alert exposure by race/ethnicity, sex, and visit context. Minority patients were more likely to receive certain alerts (e.g., SDoH, Screening) and less likely to receive others (e.g., Vaccination, Medication Stewardship), suggesting that alert logic may reinforce or mitigate existing inequities. The strong associations with visit type and comorbidity underscore the importance of clinical context in alert targeting. These findings support the need for equity audits and redesign of CDS systems to ensure fair and effective alert distribution.

Figures Figure 3, Figure 4, Figure 5, Figure 6, and Figure 7 illustrate the relative risk ratios for each alert type, with full specifications in S1 Table 4.

**Figure 3:**
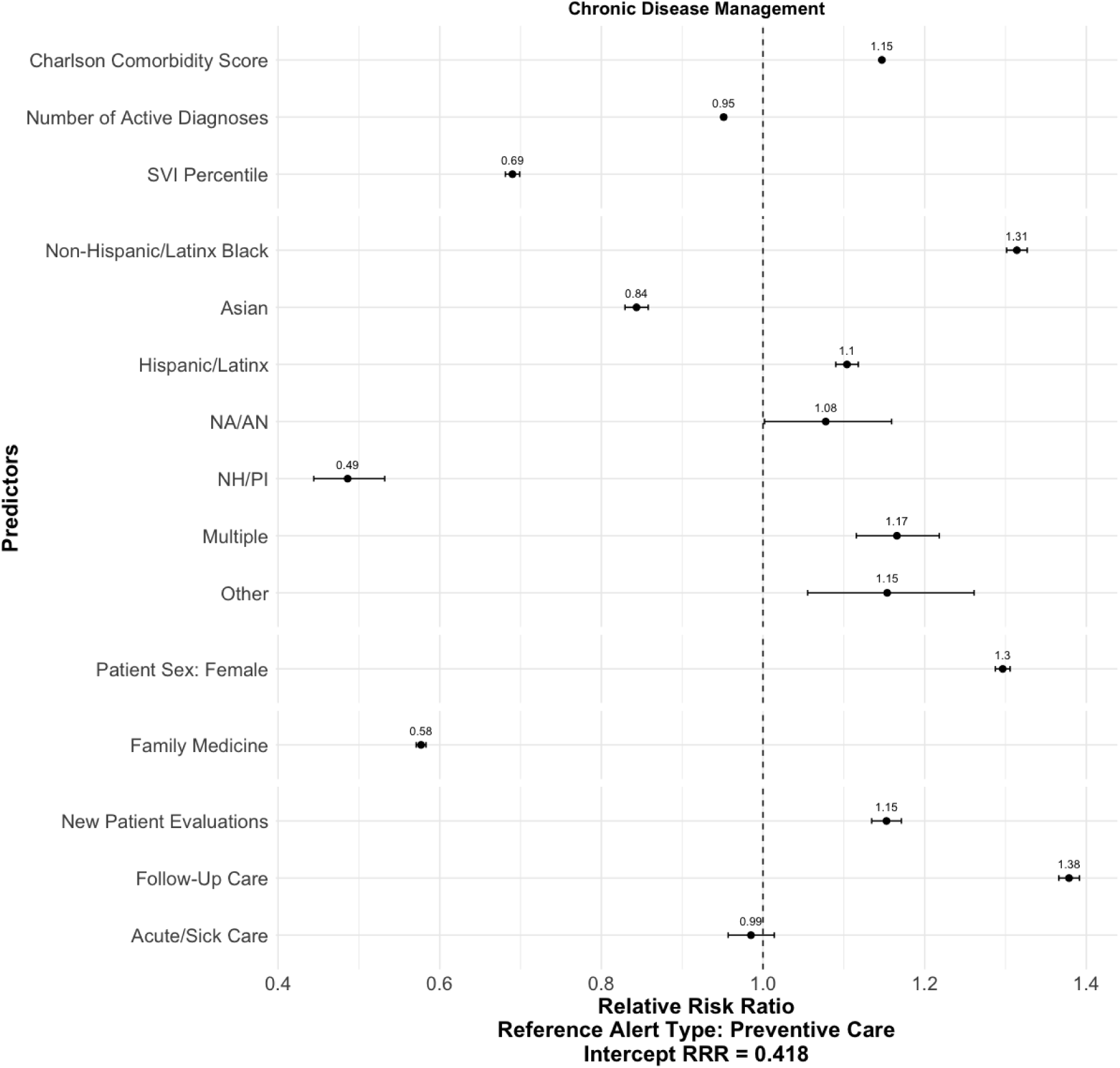
Visualization of Relative Risk Ratios from Multinomial Logistic Regression - Alerts Fired for Chronic Disease Management

**Figure 4:**
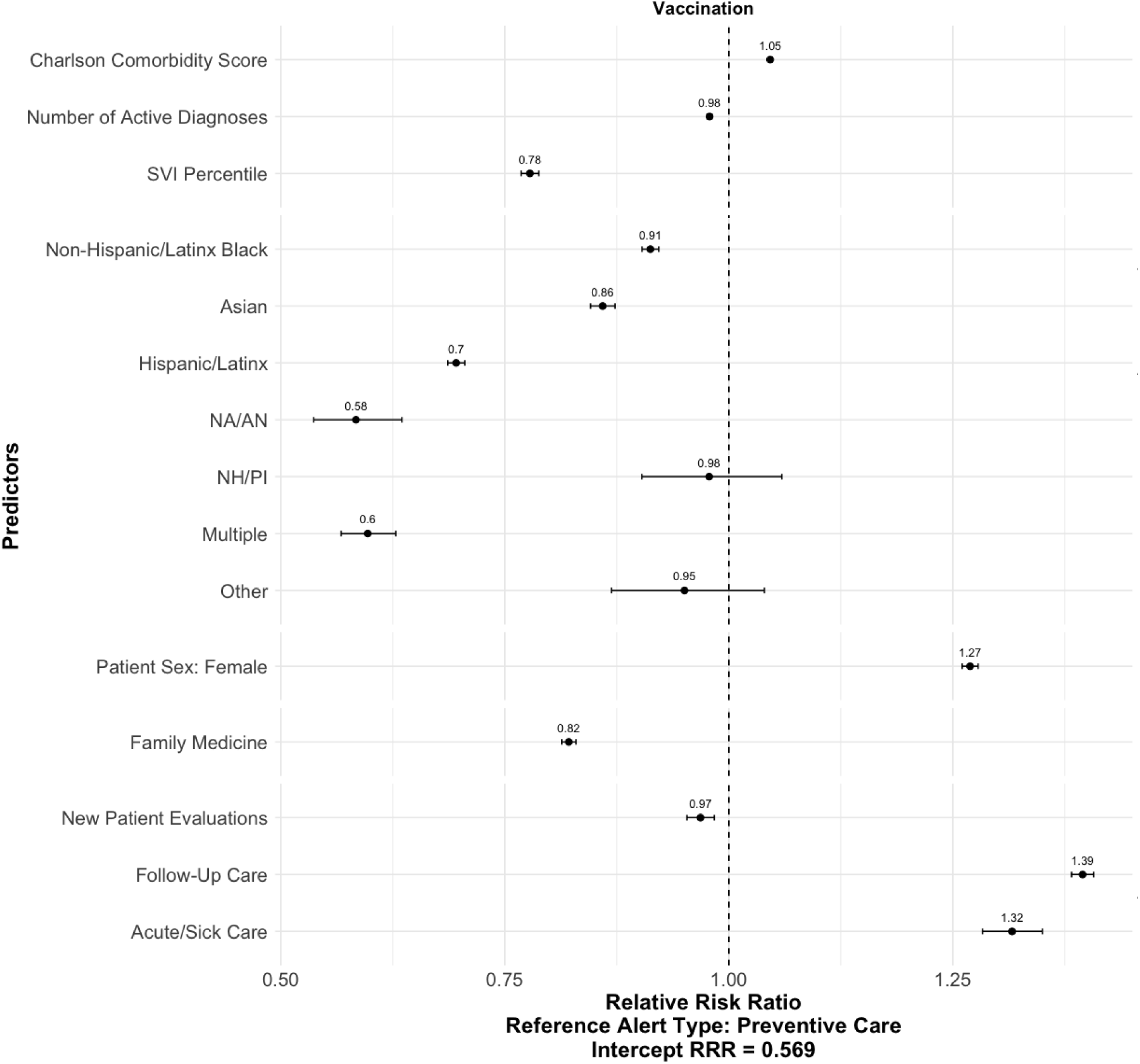
Visualization of Relative Risk Ratios from Multinomial Logistic Regression - Alerts Fired for Vaccination

**Figure 5:**
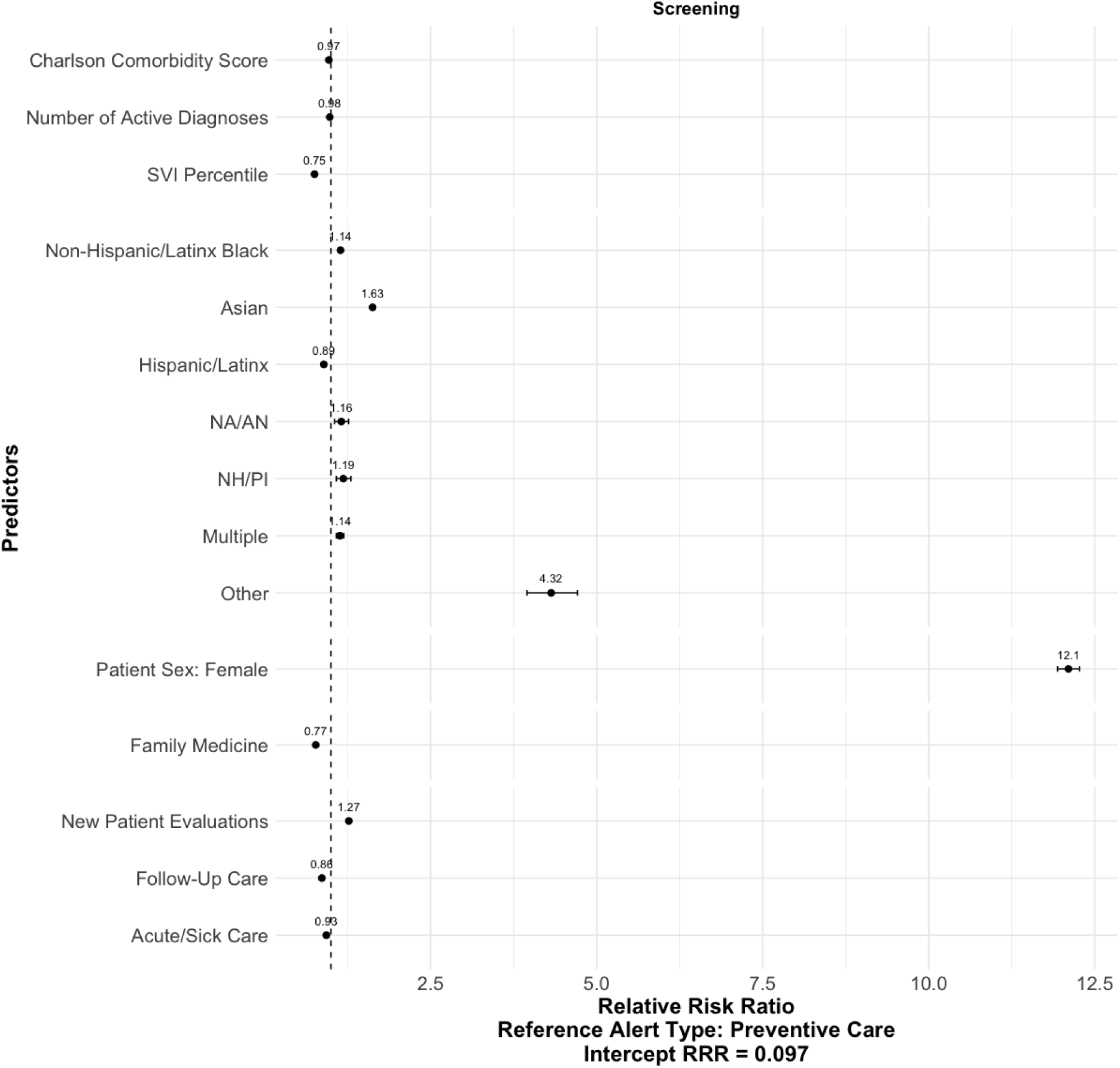
Visualization of Relative Risk Ratios from Multinomial Logistic Regression - Alerts Fired for Screening

**Figure 6:**
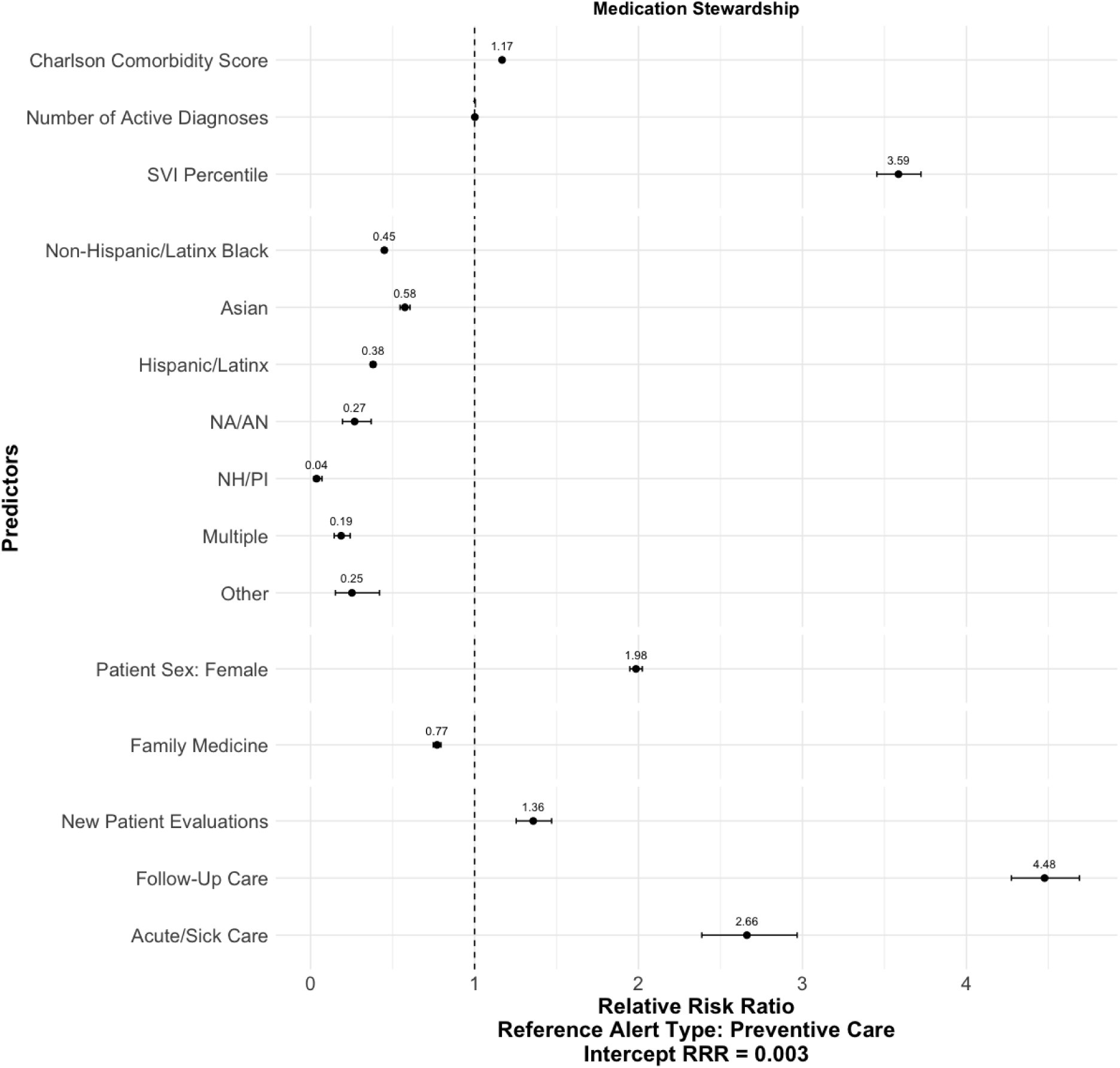
Visualization of Relative Risk Ratios from Multinomial Logistic Regression - Alerts Fired for Medication Stewardship

**Figure 7:**
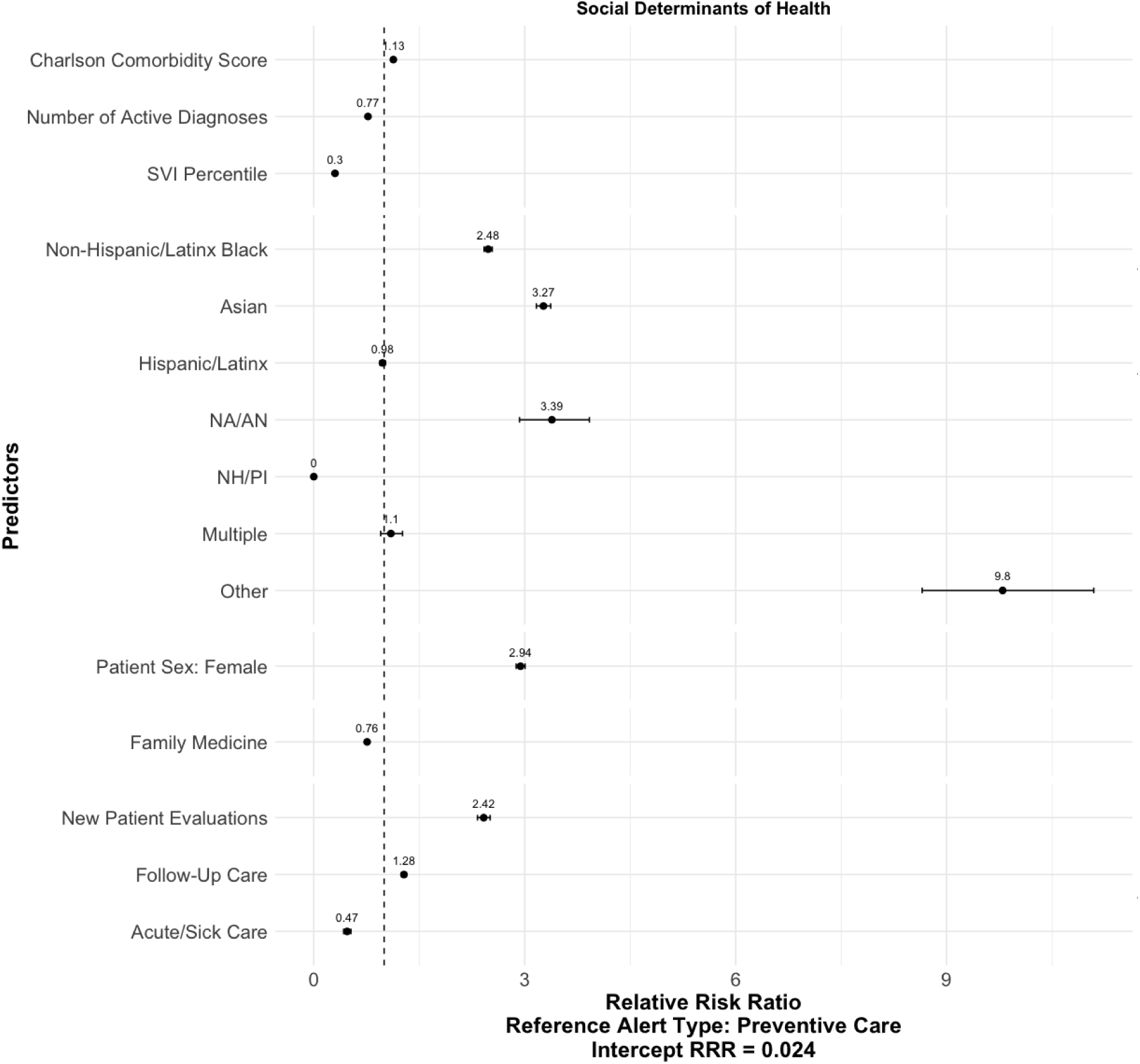
Visualization of Relative Risk Ratios from Multinomial Logistic Regression - Alerts Fired for Social Determinants of Health

### Mediation Analysis

We examined whether race/ethnicity and sex influenced the utilization of clinical decision support alerts through both direct and indirect pathways. The direct effects represent the influence of patient identity on whether alerts were acted upon, independent of alert type. The indirect effects capture the influence of race/sex on alert action via differential exposure to specific alert categories. Indirect effects were estimated using the product-of-coefficients method, with standard errors computed via the delta method. Alert-type-to-action effects (M → Y) were modeled separately to contextualize the strength of each mediation pathway.

For most racial/ethnic groups, the indirect effect of race on CDS action via alert exposure was statistically non-significant, suggesting that while alert exposure may vary by race, these differences do not reliably mediate provider action across most groups. Patients categorized as “Other” race exhibited a statistically significant negative indirect effect (β = −1.562, SE = 0.654, 95% CI [−2.844, −0.281], OR = 0.21), indicating that differential alert exposure for this group substantially reduced the likelihood of provider action. This pathway may reflect systemic gaps in CDS targeting or provider responsiveness for patients whose racial identity is not captured by standard categories. Among all alert types, only the Social Determinants of Health pathway showed a statistically significant indirect effect of race on alert action. The indirect effect via Social Determinants of Health alerts was exceptionally large and statistically significant (β = 65.060, SE = 16.742, 95% CI [32.245, 97.875], OR ≈ 1.8 × 10²⁸). This alert type had the strongest negative association with action taken (M → Y: β = −5.341, SE = 0.559), suggesting that racial disparities in exposure to Social Determinants of Health alerts may dramatically shape provider behavior. However, the magnitude of the effect and its standard error warrant cautious interpretation, as they may reflect model instability or extreme group-level differences. This suggests that disparities in exposure to Social Determinants of Health alerts may be a key driver of inequitable CDS adoption. Other alert types showed non-significant mediation effects, despite some having strong associations with provider action. These findings underscore the importance of evaluating both alert logic and exposure patterns when assessing equity in CDS systems.

For female patients, the indirect effect of sex on CDS alert follow-through via alert exposure (X → M → Y) was large, negative, and statistically significant (β = −1.184, SE = 0.311, 95% CI [−1.793, −0.575], *p* < 0.001). This suggests that women were exposed to alerts in ways that substantially reduced the likelihood of provider action. The direct effect of patient sex on alert follow-through (X → Y), independent of alert exposure, was small but significant (β = −0.063, SE = 0.030, *p* < 0.05), indicating a modest reduction in provider action even when alert type was held constant. The odds ratio for the indirect effect was 0.31 (SE = 0.095), reinforcing the magnitude of the mediation pathway. These findings suggest that alert exposure patterns among female patients may be systematically misaligned with providers’ responsiveness.

Direct and indirect effects are visualized in Figure 8 (race/ethnicity) and Figure 9 (sex). Tables of coefficients from the general structural equation model are provided in S1 Table 5 (generalized indirect effects) and S1 Table 6 (path-specific effects).

**Figure 8:**
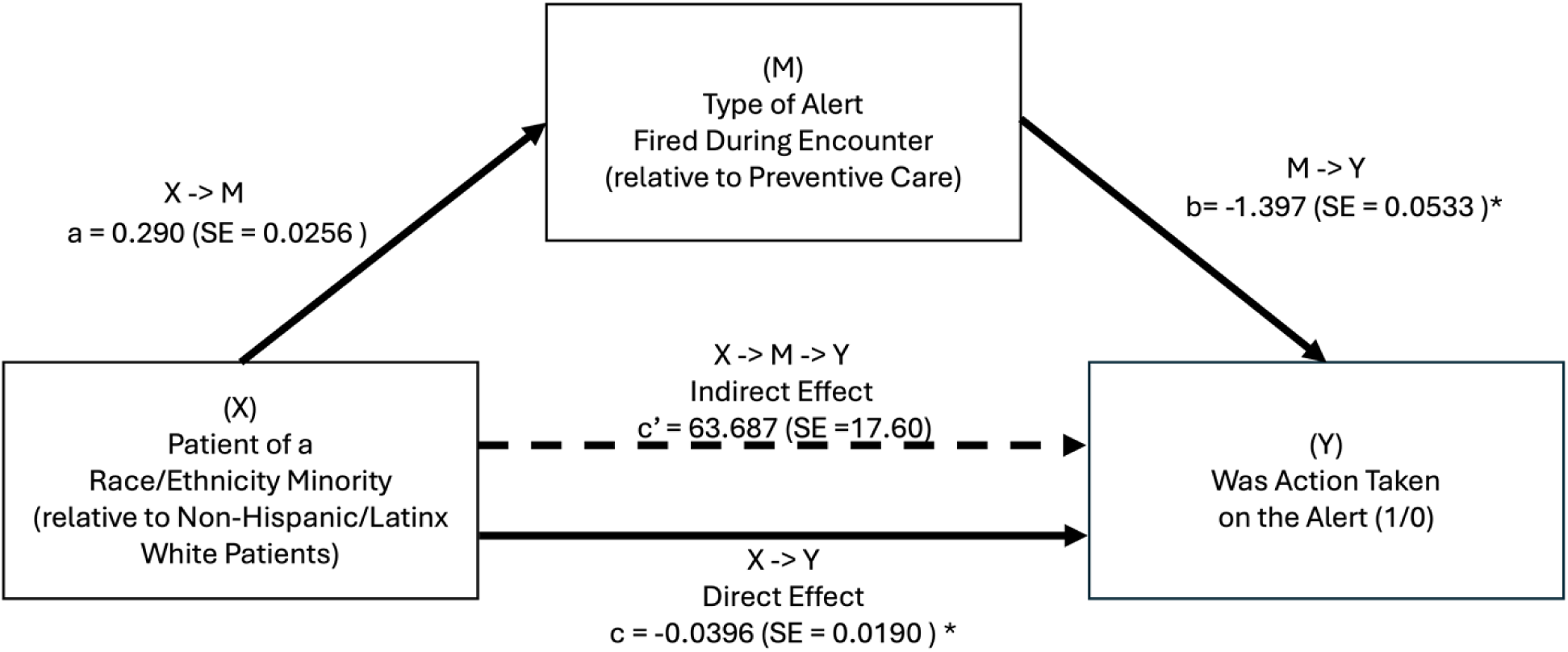
Visualization of the direct and indirect effects of a patient being of a minority race/ethnicity minority on whether action was taken through clinical decision support. The dotted arrow represents the estimated indirect effect (the effect of patient characteristics on action taken through the alert, mediated by the type of alert fired during the encounter), while the solid arrow (X->Y) represents the direct effect (the effect, independent of alert fired, of the patient sex on whether action was taken on the alert).

**Figure 9:**
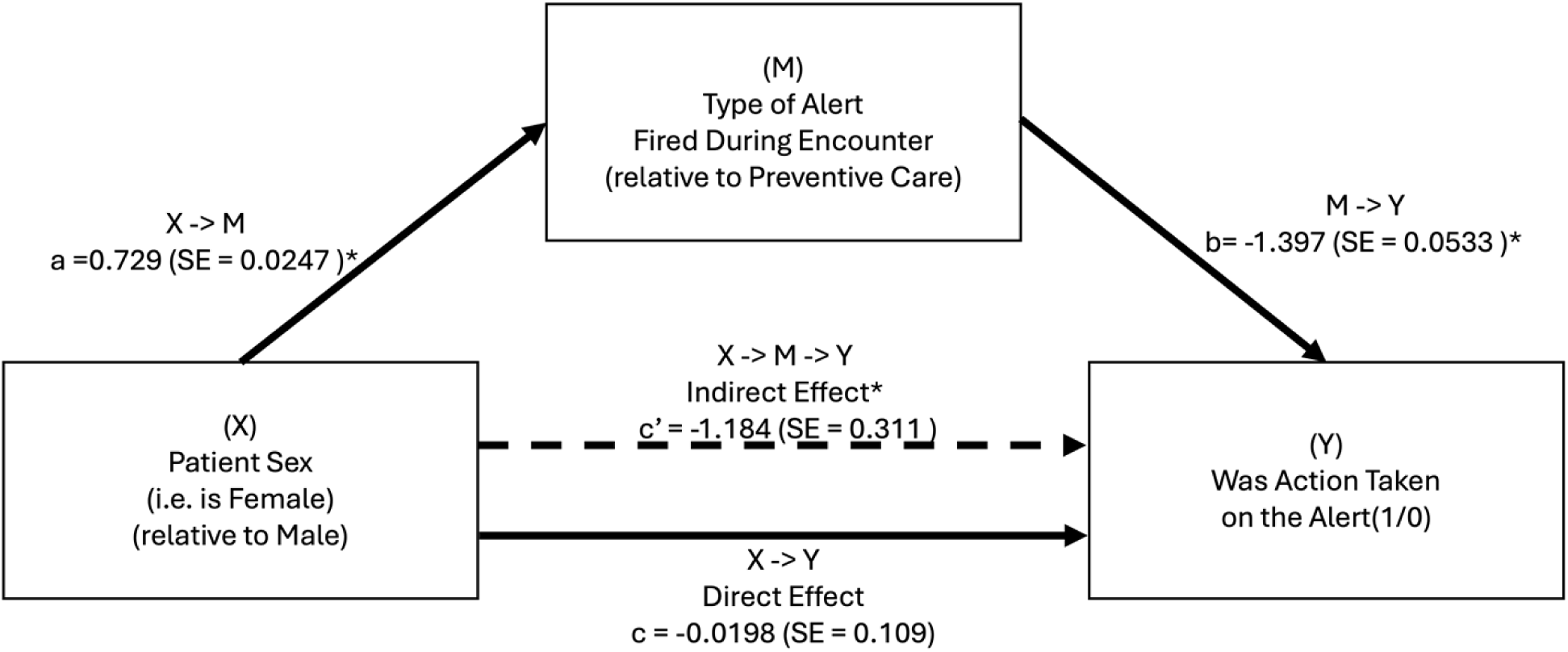
Visualization of the direct and indirect effects of a patient being female on whether action was taken through clinical decision support. The dotted arrow represents the estimated indirect effect (the effect of patient characteristics on action taken through the alert, mediated by the type of alert fired during the encounter), while the solid arrow (X->Y) represents the direct effect (the effect, independent of alert fired, of the patient sex on whether action was taken on the alert).

## DISCUSSION

### Key Results

This study examined whether provider responses to CDS alerts differed by patient race/ethnicity and sex in primary care and whether differences in alert type helped explain those patterns. By separating patient-level effects from alert- and provider-level variation, the study sought to identify potential equity gaps in CDS use within a single healthcare system and to inform more context-aware strategies for improving alert responsiveness across diverse patient populations. After accounting for alert and provider factors, patient race/ethnicity was not strongly associated with alert response, whereas visit context, alert category, and the COVID-19 period were more strongly related to whether providers acted on alerts. At the same time, alert exposure varied by patient race/ethnicity, sex, and visit context, suggesting that equity concerns arise not only in provider response but also in how alerts are generated and distributed.

Controlling for alert and provider effects showed that contextual factors, such as visit type, time of day, and the COVID-19 period, were more predictive of alert response than patient race or sex. In unadjusted models, modest race/ethnicity-based differences were observed, but these associations diminished after adjustment, suggesting that context and provider behavior may explain more of the variation. Lower response rates for certain alert categories and during busier clinical periods underscore the importance of CDS systems that are both equity-aware and workflow-sensitive.

Alert firing also differed by patient race/ethnicity, sex, and visit context. Patients from minoritized racial/ethnic groups and women were more likely to receive some alerts, such as Social Determinants of Health and Screening, and less likely to receive others, such as Vaccination and Medication Stewardship, suggesting that alert logic may contribute to uneven alert distribution. The strong associations with visit type and comorbidity further emphasize the importance of clinical context in alert targeting. These findings support the need for equity audits and redesign of CDS systems to promote fair and effective alert distribution.

The mediation analyses offer a more nuanced view of these pathways. Direct effects suggest that provider response may differ by patient identity and visit context, while indirect effects indicate differences in how alerts are assigned across groups. Together, these results suggest that even when provider response appears relatively similar after adjustment, differences in which alerts are triggered may shape the overall equity profile of CDS. Only one racial/ethnic group, patients identified as “Other,” showed a significant indirect effect of race on CDS action via alert exposure. For all other groups, indirect effects were non-significant, although several estimates, including those for NH/PI and Non-Hispanic Black patients, warrant further study in larger samples or with stratification by alert type. These patterns suggest that alert generation is an important equity target, not just provider behavior.

The negative mediation effect for female patients suggests a possible mismatch between alert logic and provider response. The alerts triggered for women may be less actionable, less trusted, or less prioritized, although this could also reflect differences in algorithm design, clinical thresholds, or documentation practices. The modest direct effect suggests that even when women receive the same alerts as men, providers may still be slightly less likely to act, but this finding should be interpreted cautiously. The magnitude and direction of these effects varied by alert type and patient group, underscoring the need for targeted follow-up.

Taken together, the results suggest that CDS equity is a sociotechnical issue. Provider decision-making, alert design, visit context, and workflow conditions all appear to contribute to differences in CDS use. Across models, alert category and encounter context were more consistently associated with CDS use than patient race or sex after adjustment. That finding argues for evaluating CDS systems at multiple points in the workflow rather than treating alert response as the sole marker of fairness. For health systems, the practical implication is that equity review should focus on both the logic that generates alerts and the conditions under which alerts are acted upon. Equity audits can identify whether particular patient groups are systematically overexposed or underexposed to specific alert types, while workflow review can identify whether providers are more likely to respond during certain visit types or periods of lower burden. These findings support a broader approach to CDS governance in which patient-level patterns, alert-level design, and provider workflow are reviewed together.

### Limitations

This study has its limitations. First, data were collected within a single academic health system, allowing the results to reflect the culture of alert response within the health system and its interaction with the enterprise health system. However, the health system comprises numerous sites across New York City, and the data were collected from 82 of them. These analyses can be replicated across healthcare systems to understand whether the patterns persist amongst other diverse healthcare populations. Because this was a retrospective cross-sectional study, selection bias may arise from restricting analyses to outpatient encounters with visible alerts and available identifiers, potentially limiting generalizability. Information bias may arise from EHR-based misclassification of race/ethnicity, sex, provider characteristics, and alert response, although the primary outcome was derived from structured alert-action data. Confounding was addressed through adjustment for patient complexity, visit context, provider characteristics, alert type, and pandemic period, all of which could influence both alert exposure and provider response.

Second, the data lake had its limitations. We did not have provider race and ethnicity, which prevents us from examining concordant patient-provider dyads. We also lack accurate representations of provider experience, despite efforts to use provider licensing dates as a proxy. We are aware of the literature on female primary care providers who face an increased documentation burden compared to their male counterparts. The study’s methods can be enhanced within healthcare systems by using HR data to examine provider characteristics associated with different care delivery to patients, thereby informing medical education and CDS design to mitigate inequitable care delivery. On the patient side, insurance information is limited in our dataset – 62.5% of the data is missing, and aggregation requirements (the greater-than-10 rule) prevented the use of meaningful categories. More comprehensive insurance information would enhance the results, enabling an examination of whether differences in coverage influence how providers make decisions about patient care.

Third, and similarly, we had limited documentation about alerts. Ideally, categorizing alerts by the taxonomy of clinical tasks associated with the alert could inform how a provider interacts with the EHR. Clinical reminders are generally categorized into test results, referrals, note-based communication, order status, patient status changes, and reminders for incomplete tasks. (21) This limitation also suggests an adverse effect of certain CDS—that they are redundant or unhelpful routes of action that could otherwise be accomplished without integrating CDS. Further categorization of clinical tasks within EHRs, not just care types, can help clarify whether alerts are the most effective mechanism for a particular task. Although additional alert metadata provided insight into how alerts were resolved (e.g., overridden, removed, canceled, or viewed), over 60% of alerts lacked this information, limiting the ability to fully characterize alert resolution or to determine whether actions were provider-initiated or automated. Further research linking these specific alerts, such as EHR audit log metadata, could provide the necessary insight. (22)

Fourth, these analyses are descriptive, not causal. Future studies that combine quantitative analysis with qualitative research—such as provider interviews—can help clarify whether disparities in CDS engagement are primarily driven by cognitive, organizational, or technological factors, thereby guiding the design of more targeted interventions. These methods can be adapted for use in randomized controlled trials and quasi-experimental designs, enabling the study of CDS utilization across various mechanisms.

### Interpreting Differences and Avoiding Race- and Sex-Based Interventions

Addressing interpersonal bias may require provider training and accountability, while mitigating algorithmic bias demands transparency in alert logic, equity audits, and inclusive design. Together, these findings underscore the need for race- and gender-sensitive CDS design, including auditing alert logic for race- and sex-based disparities in relevance and actionability, evaluating provider response patterns by sex across alert types, and incorporating race- and sex-specific clinical guidelines into CDS algorithms. Together, these findings emphasize that equity in clinical decision support is not just a matter of provider behavior — it is also a matter of system architecture.

The identified differences in CDS utilization are complex and can be understood through the framework of racialization—the processes by which racial hierarchies are created, circulated, and maintained. (23) Disparities in healthcare between racial groups reflect structural, institutional, and interpersonal forms of racism and require further research to understand and mitigate these mechanisms. (24, 25) For example, Black patients with hypertension are more likely to have uncontrolled hypertension in comparison to other racial groups, coupled with a range of complications. (26–28) However, as the results show, Black patients were no less likely than White patients to receive alerts and have those alerts acted upon.

Differences in how CDS are used can be due to ongoing legacies of racism, not an inherent or biological logic. (29) Further research could include, first, an additional collection of detailed alert covariates that can warrant systematic change by a healthcare system, and second, the use of machine learning approaches that can account for complicated nested fixed effects to analyze sources of heterogeneity in alert action. This research also enables us to investigate whether the results can be applied to non-primary care settings, such as inpatient or other ambulatory care settings.

This research raises an important ethical question: should providers and health systems strive to standardize CDS actions across all demographic groups, or is there more value in a race-conscious approach that intentionally promotes differences in CDS responses to improve the health of historically disadvantaged populations? Following outcry about racial bias in several clinical decision support tools (30), a growing medical ethics literature continues to debate how race can and should be understood in health technology (31, 32). Given the lack of progress in addressing racial inequities in American medicine, there may be value in designing CDS to be specifically race-conscious or sex-conscious to reduce inequity for marginalized groups. (33) To that end, there may be scenarios where it is ethical and ideal for providers to act more frequently on CDS for non-White patients. Future research on how and why certain types of providers and health systems respond differently to specific CDS, and on which types of CDS elicit different responses, will be critical to advancing this debate.

This paper adds to the critical literature on assessing and mitigating racial inequities in healthcare delivery by examining how providers act upon CDS differently by patient race and sex. CDS tools can be highly effective interventions, but their impact on racial equity remains underexplored. The findings from this research inform broader debates about the role of technology in promoting equitable practices among providers. Mindful design and integration of CDSs into EHRs can standardize and improve care for all populations. At the same time, prior research shows that CDS can fail if ignored (14) or if it is based on biased data and modeling. (34). By highlighting racial and gender differences in how providers act upon CDSs in practice, the findings from this research add another dimension that must be considered in the design and implementation of CDSs to promote equitable practices among providers.

Given that this paper provides evidence of limited CDS utilization for female patients, which, given more research, may be part of a larger story of female patients being marginalized in healthcare settings, these results suggest that providers must continue to work on and be aware of their own biases toward different patient populations. Our analysis cannot fully determine whether observed differences in alert engagement are driven by provider-level implicit bias, systemic factors such as EHR design and alert prioritization, or unmeasured encounter characteristics. Disparities in CDS response likely reflect a combination of these influences, as bias can operate both at the point of care and through structural mechanisms embedded in workflows and technology. While understanding these pathways is essential for future research, actionable strategies should not rely solely on assumptions about intent or direct interventions based on patient race or sex, as such approaches risk reinforcing inequities.

Instead, health systems can adopt equity-oriented strategies that address structural variability in alert use and reduce reliance on individual discretion without explicitly targeting race or sex. These include routine equity audits of CDS performance, monitoring alert response rates across demographic groups as part of quality reporting; workflow optimization to reduce alert fatigue and ensure alerts appear at clinically actionable times for all encounters; patient-centered alert logic, incorporating individualized clinical risk and social determinants rather than proxies like race or sex, which may encode historical bias; provider feedback loops using peer-benchmarked performance dashboards, reinforcing consistent CDS engagement across patient populations without race-specific directives; and cross-functional governance structures for CDS implementation, ensuring that informatics teams, clinicians, and equity officers jointly review alerts for unintended disparities before and after deployment. These universal, system-level interventions aim to standardize care delivery and mitigate inequities in CDS engagement without implementing race-based policies, which are challenging to operationalize and ethically complex.

### Broader Implications for Algorithmic Fairness and Equity Initiatives

This study contributes to the growing discourse on algorithmic fairness in healthcare by providing empirical evidence from a real-world primary care setting. As CDS becomes increasingly embedded within clinical workflows, health systems must move beyond aggregate measures of CDS effectiveness and incorporate equity metrics into the design, implementation, and evaluation of CDS. Monitoring engagement with alerts by patient demographics is an essential first step toward identifying where inequities arise and preventing downstream disparities in care delivery.

Moreover, this work underscores the need for sociotechnical approaches that examine how CDS interacts with organizational structures, clinical culture, and provider decision-making. By aligning CDS governance with health equity priorities and adopting universal strategies that minimize bias while maintaining clinical relevance, healthcare organizations can ensure that these tools fulfill their promise to improve outcomes for all patients rather than perpetuate existing inequities. In doing so, we move from asking whether CDS works to ensuring that it works equitably—an essential step toward a learning health system that promotes both quality and justice in care delivery.

## METHODOLOGY

### Ethics

This study was approved by the NYU Langone Health Institutional Review Board (IRB ID s22-008885), which waived the requirement for informed consent due to the retrospective observational design.

### Study Design

We conducted a retrospective cross-sectional observational study using EHR data and followed the Strengthening the Reporting of Observational Studies in Epidemiology (STROBE) guidelines for reporting observational research. (35) Although CDS encompasses multiple modalities, including order sets, documentation templates, and predictive tools, this study limits its scope to alert-based CDS because they are easy to track in EHR data. We focused on alerts designed to prompt guideline-concordant actions during outpatient encounters, as these represent the most common and influential type of CDS in primary care.

### Setting

The study was conducted within a large academic healthcare system in New York City, serving a diverse urban population across multiple boroughs. The system comprises primary care, family medicine, and geriatric practices that utilize a single, integrated EHR platform. Supplemental data on providers’ sex were pulled from NPI databases. Alert-firing data were extracted from the healthcare system’s data lake, built on a Hadoop distributed file system, and relevant data were queried using MySQL databases. We restricted the sample to practices in which responding to alerts was not tied to provider or clinic incentives, thereby isolating alert use independent of financial requirements (e.g., federally qualified health centers, which may mandate specific alert response rates (36)

### Study Period

We analyzed data from two non-overlapping 12-month periods: the pre-COVID-19 pandemic period, from July 1, 2018, to June 30, 2019, and a symmetric post-acute COVID-19 period, from July 1, 2021, to June 30, 2022. These periods were selected to control for changes in CDS alert utilization before and after the COVID-19 pandemic and to account for pandemic-related workflow disruptions. We included all eligible encounters within the prespecified study periods, thereby capturing the full available EHR sample for this setting.

### Inclusion and Exclusion

Eligible encounters were those in primary care, family medicine, or geriatric care practices within the health system, involving patients aged 18 years or older who had at least one alert firing during their encounter. We excluded encounters with missing patient identifiers and encounters in specialty clinics, urgent care, and inpatient settings. We restricted analyses to alerts explicitly displayed to providers during the encounter, ensuring that the outcome reflected alerts visible within the clinical workflow and available for provider action.

### Outcome and Mediator Variable: Clinical Decision Support Alerts and Responsiveness

We examined alert-based CDS prompts that encourage evidence-based care for the management of preventive and chronic diseases. Ninety-six alerts were categorized into six content categories based on a taxonomy provided in the EHR system and the alert name. Alert categorizations are listed in S1 Table 7. The primary outcome variable was a binary indicator reflecting whether the provider took a meaningful clinical action in response to an alert. Specifically, it was coded as a 1 if the advisory triggered an action (e.g., opening an order set, placing an order, documenting a diagnosis) other than an acknowledgment, deferral, or dismissal. If no such action was taken, the variable was coded as a 0. This measure captured whether the alert prompted the provider to engage with the advisory in a clinically substantive way. We excluded alerts related to administrative support and research from the analysis of whether an alert was responded to but included them in the count of alerts a provider could act upon during a patient encounter.

### Exposure Variables

The primary exposures were the patient’s self-identified race, recorded in categories based on U.S. census categorizations (37), and the patient’s sex. Covariates at the patient level included age, the Social Vulnerability Index (38), the number of active problems in the patient’s medical records, and the Charlson Comorbidity Index. (39) Encounter-level covariates included the visit category, whether the encounter was before or during the COVID-19 pandemic, the day of the week, the hour of the visit, and the number of alerts shown to the provider during the encounter. Provider-level variables available were provider type (physician, nurse practitioner (NP), or physician assistant (PA)), department (Internal Medicine or Family Medicine), and sex (male or female). Covariates were selected a priori based on clinical relevance and data availability in the EHR, focusing on factors that could influence alert generation or provider response, including patient complexity, encounter context, provider characteristics, and pandemic timing.

### Statistical Analysis

Analyses were conducted using Stata/SE 18.5.

Descriptive statistics summarized patient, provider, alert, and encounter characteristics. Continuous variables were summarized as means (standard deviations) and medians (minimum, maximum), and categorical variables as frequencies (%). Continuous variables were modeled continuously to preserve information.

We conducted a series of regression analyses to examine the relationship between patient demographic characteristics and alert-related outcomes. First, logistic regression models were used to estimate the likelihood that an alert would result in clinical action, incorporating both patient-level and encounter-level variables. To account for heterogeneity in alert behavior and repeated measures within providers, we included fixed effects for alert ID and clustered standard errors at the provider level. We estimated alternative model specifications—with and without alert fixed effects, provider clustering, and unknown values for patient race and ethnicity, as well as sex—to assess how these adjustments influenced estimates of demographic associations. Regressions were estimated using a Poisson pseudo-maximum-likelihood estimator with high-dimensional fixed effects. We chose this approach because it is robust to separation issues and allows estimation with many fixed effects and a large number of zeros, while still enabling consistent estimates of conditional odds ratios. (40) Regression coefficients were exponentiated to yield adjusted risk ratios (ARRs) for interpretability.

A provider’s response to an alert may depend on the type of alert they see, and various patient characteristics may influence the type of alert that is fired for them. Therefore, we conducted a multinomial logistic regression analysis to examine the relationship between the firing of an alert in the six identified categories, patient characteristics, and the department where a patient had their primary care encounter. To estimate the mediating effect of the alert type on the relationship between a patient’s race and sex and a provider’s response to an alert, we specified a generalized structural equation model. This model simultaneously estimated both the multinomial logistic regression predicting alert type based on patient and visit characteristics, and the Poisson regressions with a log link, modeling the probability of the alert action taken conditional on the alert type, patient and visit characteristics, and available provider characteristics. Indirect and direct effects were calculated using nonlinear combinations of parameters, as per Stata’s nlcom-based mediation estimation framework, thereby decomposing the total effect of patient race and sex on alert response into components mediated by alert type and those acting directly. These analyses would differentiate between the direct effect of patient race and sex on a provider’s response to an alert and the indirect effect of alert type. (41, 42)

## ACKNOWLEDGMENTS

We thank Hao Zhang for initial guidance on data pulling. We thank Saul Blecker for early feedback on writing the manuscript. We thank audiences at the Society of General Internal Medicine Annual Conference and AcademyHealth’s Annual Research Meeting in 2023 for their feedback.

## DATA AVAILABILITY

Deidentified data are available upon request.

## AUTHOR CONTRIBUTIONS (CRediT TAXONOMY)

| Contributor Role | Role Definition | Author |
| --- | --- | --- |
| **Conceptualization** | Ideas; formulation or evolution of overarching research goals and aims. | RVNV, SIR |
| **Data Curation** | Management activities to annotate (produce metadata), scrub data and maintain research data (including software code, where it is necessary for interpreting the data itself) for initial use and later reuse. | RVNV |
| **Formal Analysis** | Application of statistical, mathematical, computational, or other formal techniques to analyze or synthesize study data. | RVNV, SIR, SAJ |
| **Funding Acquisition** | Acquisition of the financial support for the project leading to this publication. | RVNV, SIR |
| **Investigation** | Conducting a research and investigation process, specifically performing the experiments, or data/evidence collection. | RVNV |
| **Methodology** | Development or design of methodology; creation of models | RVNV, SAJ |
| **Project Administration** | Management and coordination responsibility for the research activity planning and execution. | RVNV |
| **Resources** | Provision of study materials, reagents, materials, patients, laboratory samples, animals, instrumentation, computing resources, or other analysis tools. | SIR |
| **Software** | Programming, software development; designing computer programs; implementation of the computer code and supporting algorithms; testing of existing code components. | RVNV |
| **Supervision** | Oversight and leadership responsibility for the research activity planning and execution, including mentorship external to the core team. | SIR |
| **Validation** | Verification, whether as a part of the activity or separate, of the overall replication/reproducibility of results/experiments and other research outputs. | N/A |
| **Visualization** | Preparation, creation and/or presentation of the published work, specifically visualization/data presentation. | RVNV |
| **Writing – Original Draft Preparation** | Creation and/or presentation of the published work, specifically writing the initial draft (including substantive translation). | RVNV, KO, SAJ, SIR |
| **Writing – Review & Editing** | Preparation, creation and/or presentation of the published work by those from the original research group, specifically critical review, commentary or revision – including pre- or post-publication stages. | RVNV, KO, SAJ, SIR |

## SUPPORTING INFORMATION CAPTIONS

S1 Table 1: Comparison of Regressions Including Unknown Patient Race/Ethnicity, Other/Unknown Patient Sex, Clustering Errors by Provider, and Fixed Effects of Alert Type

S1 Table 2: Logistic Regression Coefficients – Adjusted Model

S1 Table 3: Logistic Regression Coefficients – Unadjusted Model

S1 Table 4: Multinomial Logistic Regression Results

S1 Table 5: Full Estimates from General Structural Equation Model

S1 Table 6: Estimates of Aggregated Indirect Paths

S1 Table 7: Alert Categorization and Frequency of Firing in Study Period

